# Gut microbiome diversity is an independent predictor of survival in cervical cancer patients receiving chemoradiation

**DOI:** 10.1101/2020.05.18.20105809

**Authors:** Travis T. Sims, Molly B. El Alam, Tatiana V. Karpinets, Stephanie Dorta-Estremera, Venkatesh L. Hegde, Sita Nookala, Kyoko Yoshida-Court, Xiaogang Wu, Greyson W. G. Biegert, Andrea Y. Delgado Medrano, Travis Solley, Mustapha Ahmed-Kaddar, Bhavana V. Chapman, K. Jagannadha Sastry, Melissa P. Mezzari, Joseph F. Petrosino, Lilie L. Lin, Lois Ramondetta, Anuja Jhingran, Kathleen M. Schmeler, Nadim J Ajami, Jennifer Wargo, Lauren E. Colbert, Ann H. Klopp

## Abstract

Diversity of the gut microbiome is associated with higher response rates for cancer patients receiving immunotherapy but has not been investigated in patients receiving radiation therapy. Additionally, current studies investigating the gut microbiome and outcomes in cancer patients may not adjusted for established risk factors. Here, we sought to determine if diversity and composition of the gut microbiome was independently associated with survival in cervical cancer patients receiving chemoradiation. Our study demonstrates that the diversity of gut microbiota is associated with a favorable response to chemoradiation. Additionally, compositional variation among patients correlated with short term and long-term survival. Short term survivor fecal samples were significantly enriched in *Porphyromonas, Porphyromonadaceae*, and *Dialister*, whereas long term survivor samples were significantly enriched in *Escherichia Shigella, Enterobacteriaceae*, and *Enterobacteriales*. Moreover, analysis of immune cells from cervical tumor brush samples by flow cytometry revealed that patients with a high microbiome diversity had increased tumor infiltration of CD4+ lymphocytes as well as activated subsets of CD4 cells expressing ki67+ and CD69+ over the course of radiation therapy. The modulation of gut microbiota before chemoradiation might provide an alternative way to enhance treatment efficacy and improve treatment outcomes in cervical cancer patients.

## MAIN

Cervical cancer continues to be one of the leading causes of cancer-associated mortality globally^1^. More than 500,000 new cases of invasive cervical cancer will be diagnosed worldwide in 2020, resulting in over 300,000 deaths^2^. Multimodality therapy consisting of definitive chemoradiation (CRT) comprising external-beam radiotherapy (EBRT) followed by intracavitary brachytherapy with concurrent systemic chemotherapy continues to be the standard of care in clinical practice for locally advanced disease^3^.

The fecal or gut microbiome, a diverse community of bacteria, archaea, fungi, protozoa, and viruses, is thought to influence host immunity by modulating multiple immunologic pathways, thus impacting health and diseases^4-6^. The diversity of the gut microbiome is defined as the number and relative abundance distribution of these distinct types of microorganisms colonizing within the gut^7^. Studies have suggested that dysbiosis of the gut microbiome confers a predisposition to certain malignancies and influences the body’s response to a variety of cancer therapies, including chemotherapy, radiotherapy, and immunotherapy^6,8–11^. For example, melanoma patients are more likely to have a favorable response to immune checkpoint blockade and exhibit improved systemic and antitumor immunity if they have a more diverse intestinal microbiome^11^.

Radiotherapy promotes the activation of T cells directed against tumor antigens^12-15^. In combination with immunotherapy, radiotherapy can maximize the antitumor immune response and promote durable disease control^16,17^. We theorize that the gut microbiota may modulate radioresponse through immunologic mechanisms^14,18^. Studies investigating the gut microbiome and outcomes in cancer patients often do not adjust for confounding patient and tumor characteristics. To assess this, we sought to identify independent gut microbial risk factors in cervical cancer (CC) patients receiving chemoradiation (CRT) and to evaluate their impact on survival. We hypothesize that gut microbial differences may affect clinical outcomes in patients with cervical cancer

## RESULTS

### Patient Characteristics

A total of 55 patients with a mean age of 47 years (range, 29-72 years) volunteered to participate in this study. The patients received standard treatment for cervical cancer with 5 weeks of EBRT and weekly cisplatin. After completion of EBRT, patients received brachytherapy. For evaluation of treatment response, patients underwent magnetic resonance imaging (MRI) at baseline and week 5 and positron emission tomography (PET)/computed tomography (CT) 3 months after treatment completion (Fig. 1a). Most patients had stage IIB disease (51%) and squamous histology (78%). Their clinicopathologic data are summarized in Supplementary Table 1. We staged cervical cancer using the 2014 International Federation of Gynecology and Obstetrics staging system. The median cervical tumor size according to MRI was 5.4 cm (range, 1.2-11.5 cm). Thirty patients (55%) had lymph node involvement according to imaging studies. We first analyzed the bacterial 16S rDNA (16Sv4) fecal microbiota at baseline with respect to disease histology, grade, and stage. We found that the baseline α-diversity (within tumor samples) and β-diversity (between samples) of the fecal microbiome in the cervical cancer patients did not differ according to histology, grade, or stage (*P* > 0.05) (Supplementary Fig. 1a-d).

**Fig 1.**
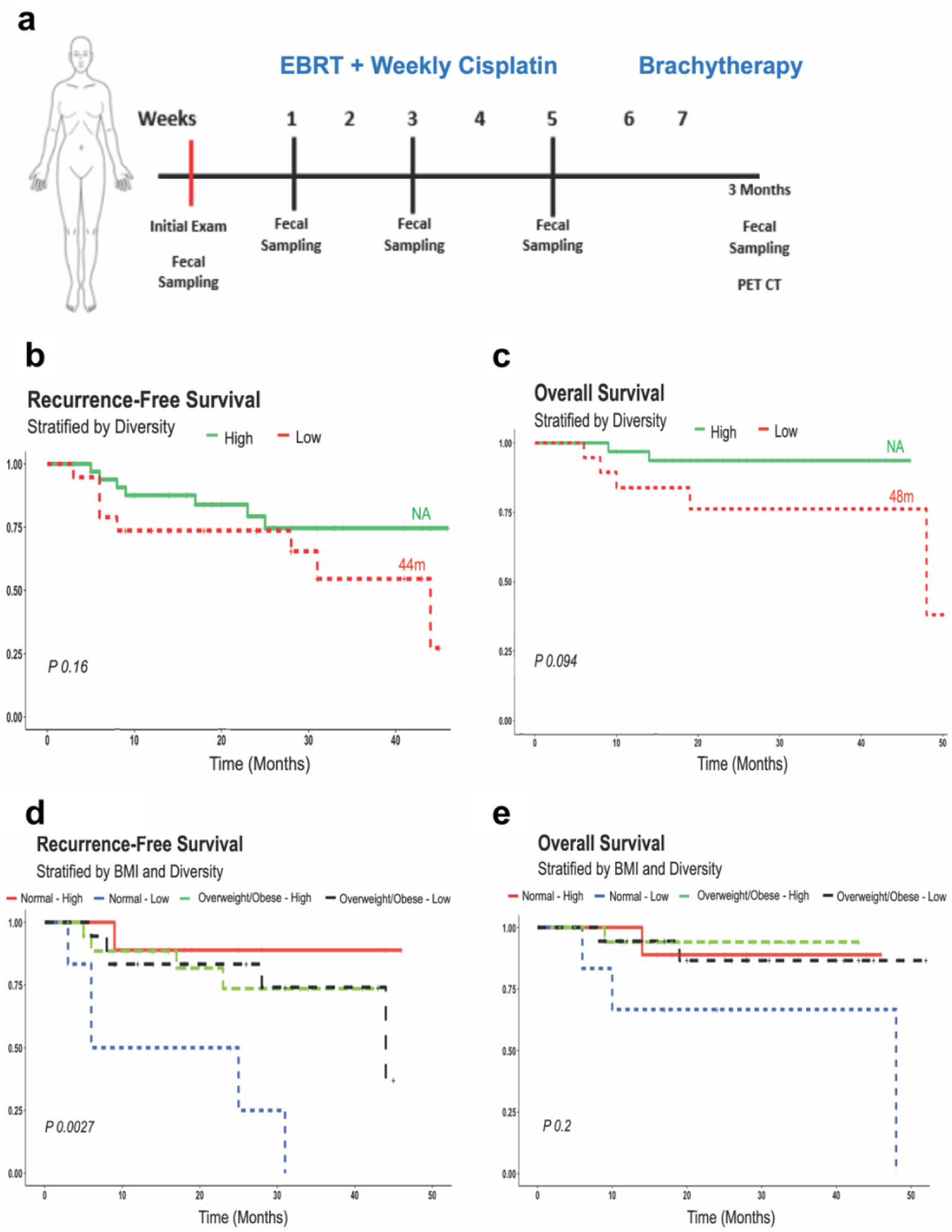
Relationship between gut diversity and BMI. (A) Schema of the sample collection, treatment, and analyses used in the present study. Kaplan-Meier curves for (B) recurrence free survival, (C) overall survival stratified by high and low gut diversity. Kaplan-Meier curves for (D) recurrence free survival, (E) overall survival stratified by BMI and gut diversity. Cases represent patients.

### Univariate and multivariate analysis of factors affecting recurrence free survival (RFS) and overall survival (OS)

In the univariate Cox proportional hazard regression model predicting RFS, 3 covariates showed P ≤ 0.2. As shown in Table 1, univariate analysis identified older age (Hazard Ratio (HR) of 0.93 (95% CI = 0.87-0.98, P = 0.0096)), Shannon diversity index (SDI) (HR of 0.51 (95% CI = 0.23-1.1, P = 0.087)) and body mass index (BMI) (HR of 0.92 (95% CI = 0.84-1, P = 0.096)) as risk factors for RFS. Multivariate survival analyses identified BMI and SDI as independent prognostic factors for RFS with a HR of 0.87 (95% CI = 0.77-0.98, P = 0.02) and 0.36 (95% CI = 0.15-0.84, P = 0.018) respectively. As shown in Table 2, univariate analysis identified SDI (HR of 0.34 (95% CI = 0.1-1.1, P = 0.08) and BMI (HR of 0.83 (95% CI = 0.69-1, P = 0.055)) as risk factors for OS. For OS, multivariate survival analyses again identified BMI and SDI as independent prognostic factors with a HR of 0.78 (95% CI = 0.623-0.97, P = 0.025) and 0.19 (95% CI = 0.043-0.83, P = 0.028) respectively.

**Table 1.** Univariate and multivariate Cox regression analysis for recurrence-free survival.

| Characteristics | Univariate model |  | Multivariate model |  |
| --- | --- | --- | --- | --- |
|  | HR (95% CI) | *P* value | HR (95% CI) | *P* value |
| **Age** | 0.93 (0.87-0.98) | 0.0096 | 0.93* (0.88-0.99) | 0.03‡ |
| **BMI (kg/m2)** | 0.92 (0.84-1) | 0.096 | 0.87* (0.77-0.98) | 0.02‡ |
| Normal (18.5 to 24.9) | 1 (reference) |  | — | — |
| Overweight (25 to 29.9) | 0.81(0.26-2.53) | 0.715 | — | — |
| Obese (30 or more) | 0.47(0.13-1.67) | 0.240 | — | — |
| **Race/Ethnicity** |  |  |  |  |
| Asian | 1 (reference) |  | — | — |
| Black | 0.37(0.02-5.90) | 0.479 | — | — |
| Hispanic | 0.39 (0.05-3.21) | 0.382 | — | — |
| White | 0.39 (0.05-3.31) | 0.390 | — | — |
| Other | 4.1309E-08(-inf - +inf) | 0.998 | — | — |
| **Stage** |  |  |  |  |
| I | 1 (reference) |  | — | — |
| II | 1.50 (0.31-7.34) | 0.615 | — | — |
| III | 3.99 (0.80-20.01) | 0.091 | — | — |
| IV | 2.54 (0.23-28.12) | 0.447 | — | — |
| **Grade** |  |  |  |  |
| Well | 1 (reference) |  |  |  |
| Moderate | 55297546(-inf - +inf) | 0.998 | — | — |
| Poor | 97336741.9(-inf - +inf) | 0.998 | — | — |
| Unknown | 76285161.4(-inf - +inf) | 0.998 | — | — |
| **Histology** |  |  |  |  |
| Squamous | 1 (reference) |  | — | — |
| Adenocarcinoma/Adenosquamous | 1.06(0.34-3.34) | 0.918 | — | — |
| **Node Level on PET** |  |  |  |  |
| Common Iliac | 1 (reference) |  | — | — |
| External Iliac | 1.33 (0.35-4.95) | 0.675 | — | — |
| Internal Iliac | 0.67 (0.07-6.89) | 0.736 | — | — |
|  | HR (95% CI) | *P* value | HR (95% CI) | *P* value |
| Para-Aortic | 1.31 (0.14-12.55) | 0.818 | — | — |
| None | 0.34 (0.06-2.09) | 0.245 | — | — |
| **Max Tumor Dimension on MRI** | 1.3 (1-1.8) | 0.042 | — | — |
| **Smoking status** |  |  |  |  |
| Current | 1 (reference) |  | — | — |
| Former | 0.91 (0.10-7.84) | 0.934 | — | — |
| Never | 0.89(0.11-7.17) | 0.909 | — | — |
| **Antibiotic Use** |  |  |  |  |
| No | 1 (reference) |  | — | — |
| Yes | 78371200.7 (-inf - +inf) | 0.998 | — | — |
| **Brachytherapy** |  |  |  |  |
| HDR | 1 (reference) |  | — | — |
| PDR | 1.41 (0.48-4.149) | 0.532 | — | — |
| **Baseline Gut Alpha Diversity** |  |  |  |  |
| Observed OTU | 0.99 (0.97-1) | 0.21 | — | — |
| Shannon | 0.51 (0.23-1.1) | 0.087 | 0.36* (0.15-0.84) | 0.018‡ |
| Simpson | 0.025 (0.000036-1.7) | 0.087 | — | — |
| Inverse Simpson | 0.93 (0.84-1) | 0.11 | — | — |
| Fisher | 0.95 (0.88-1) | 0.23 | — | — |
| Camargo | 13 (0.14-1300) | 0.27 | — | — |
| Pielou | 0.02 (0.00026-1.6) | 0.081 | — | — |

*CI*, Confidence interval; *HR*, hazard ratio.
\*Significant hazard ratios.
‡Significant *P* value.

**Table 2.** Univariate and multivariate Cox regression analysis for overall survival.

| Characteristics | Univariate model |  | Multivariate model |  |
| --- | --- | --- | --- | --- |
|  | HR (95% CI) | P value | HR (95% CI) | P value |
| **Age** | 0.95 (0.87-1) | 0.23 | — | — |
| **BMI (kg/m2)** | 0.83 (0.69-1) | 0.055 | 0.78* (0.623-0.97) | 0.025‡ |
| Normal (18.5 to 24.9) | 1 (reference) |  | — | — |
| Overweight (25 to 29.9) | 0.23(0.08-2.32) | 0.323 | — | — |
| Obese (30 or more) | 0.42 (0.06-4.56) | 0.19 | — | — |
| **Race/Ethnicity** |  |  |  |  |
| Asian | 1 (reference) |  | — | — |
| Black | 4.46E-09 (-inf - +inf) | 0.999 | — | — |
| Hispanic | 0.23(0.02-2.22) | 0.204 | — | — |
| White | 0.17 (0.02-1.90) | 0.151 | — | — |
| Other | 4.48E-09 (-inf - +inf) | 0.999 | — | — |
| **Stage** |  |  |  |  |
| I | 1 (reference) |  | — | — |
| II | 1.19 (0.12-11.43) | 0.881 | — | — |
| III | 1.49 (0.09-23.93) | 0.776 | — | — |
| IV | 5.13 (0.32-82.34) | 0.248 | — | — |
| **Grade** |  |  |  |  |
| Well | 1 (reference) |  | — | — |
| Moderate | 116103697.1 (-inf - +inf) | 0.999 | — | — |
| Poor | 46065187.92(-inf - +inf) | 0.999 | — | — |
| Unknown | 149251105.9(-inf - +inf) | 0.999 | — | — |
| **Histology** |  |  |  |  |
| Squamous | 1 (reference) |  | — | — |
| Adenocarcinoma/Adenosquamous | 3.40 (0.69-16.90) | 0.134 | — | — |
| **Node Level on PET** |  |  |  |  |
| Common Iliac | 1 (reference) |  | — | — |
| External Iliac | 1.099 (0.09-12.86) | 0.306 | — | — |
| Internal Iliac | 4.83 (0.24-98.040) | 0.999 | — | — |
| Para-Aortic | 5.9333E-08 (-inf - +inf) | 0.354 | — | — |
|  | HR (95% CI) | *P* value | HR (95% CI) | *P* value |
| None | 3.34(0.26-42.69) | 0.940 | — | — |
| **Max Tumor Dimension on MRI** | 1.2 (0.77-1.8) | 1.2 | — | — |
| **Smoking status** |  |  |  |  |
| Current | 1 (reference) |  | — | — |
| Former | 106318829.2 (-inf - +inf) | 0.999 | — | — |
| Never | 61091037.65 (-inf - +inf) | 0.999 | — | — |
| **Antibiotic Use** |  |  |  |  |
| No | 1 (reference) |  | — | — |
| Yes | 0.53 (0.06-4.56) | 0.564 | — | — |
| **Brachytherapy** |  |  |  |  |
| HDR | 1 (reference) |  | — | — |
| PDR | 0.89 (-1.61-1.39) | 0.884 | — | — |
| **Baseline Gut Alpha Diversity** |  |  |  |  |
| Observed OTU | 0.98 (0.95-1) | 0.14 | — | — |
| Shannon | 0.34 (0.1-1.1) | 0.08 | 0.19* (0.043-0.83) | 0.028‡ |
| Simpson | 0.0059 (1.2e-05-2.9) | 0.1 | — | — |
| Inverse Simpson | 0.85 (0.7-1) | 0.13 | — | — |
| Fisher | 0.91 (0.79-1) | 0.15 | — | — |
| Camargo | 2200 (0.84-5800000) | 0.055 | — | — |
| Pielou | 0.0036 (5e-06-2.5) | 0.093 | — | — |

*CI*, Confidence interval; *HR*, hazard ratio.
\*Significant hazard ratios.
‡Significant *P* value.

### Baseline Gut Microbiota Diversity is Associated with Favorable Responses

During the median follow-up period of 24.5 months, 7 patients died; all patients (12.7% of the total study population) died of disease (DOD). Figure 1 shows the Kaplan-Meier curves for RFS and OS. Given our univariate and multivariate analyses performed by Cox proportional hazard model identified SDI as an independent predictor for RFS and OS, we first tested the relationship between gut diversity and RFS and OS in our cohort by stratifying patients based on high and low SDI. We stratified the patients by SDI as high-diversity versus low-diversity groups based on the cutoff value of SDI (2.69) calculated by receiver operating characteristic curve (ROC). We demonstrate that patients with high fecal alpha diversity at baseline showed a trend toward prolonged RFS and OS when compared to those with low diversity (P = 0.16 and 0.094, respectively) (Fig. 1a,b). Next, because our univariate and multivariate analyses performed by Cox proportional hazard model also identified BMI as an independent predictor for RFS and OS, we tested the relationship between diversity and RFS and OS in our cohort by stratifying patients based on high and low Shannon diversity metric and normal or high BMI. As shown in Figure 1d,e, when BMI and gut diversity are stratified for at baseline, patients with normal BMI and higher SDI had a longer median RFS duration (P = 0.0027) (Fig. 1d). OS (Fig. 1e) was longer for patients with normal BMI and higher gut diversity (P = 0.2).

**Fig. 2.**
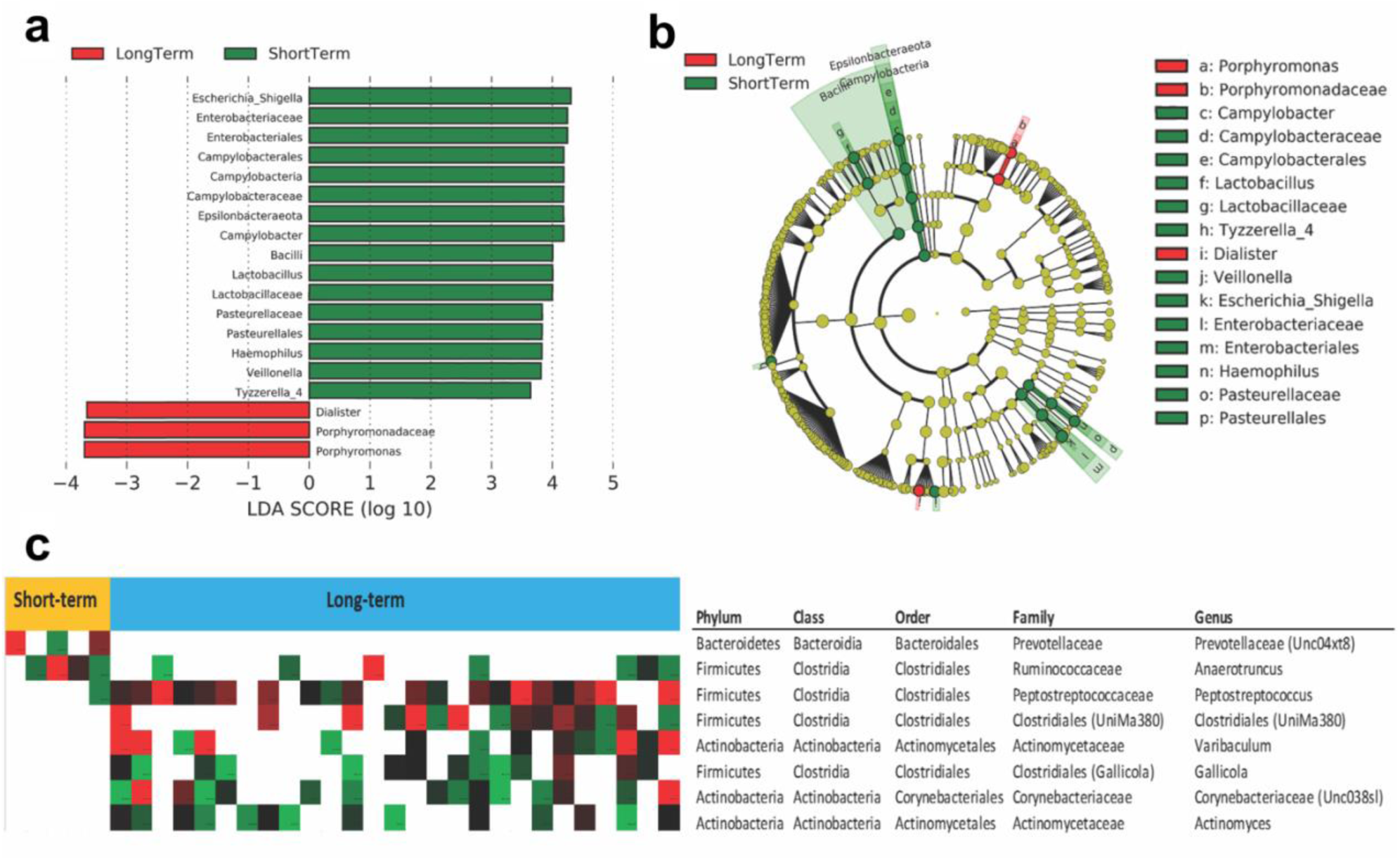
Compositional differences of the gut microbiome in short and long term survivors. (A) The different abundance of bacterial genus between the two groups were identified by LEfSe. It was significantly different when alpha value of the factorial Kruskal–Wallis test was <0.05 and the logarithmic LDA score was >3.0. The left histogram showed the LDA scores of genera differentially abundant between the two groups. The taxonomy was listed, followed by its core group. Putative species (Specific OTUs) identified as significantly more enriched/depleted (Fisher/Wilcoxon test p value < 0.05) in patients with short-term vs long-term in baseline samples. (B) Cladogram representation of the significantly different taxa features from phylum (inner circle) to genus (outer circle) (C) The right heatmap showed the relative abundance of specific bacteria by phylum, class, order, family and genus between short-term and long-term survivors.

### Compositional Difference in Gut Microbiome in Response to chemoradiation

To further investigate whether the composition of gut microbiome was associated with response to CRT, we used Linear discriminant analysis (LDA) Effect Size analysis to identify bacterial genera that were differentially enriched in short term and long term cervical cancer patients (P < 0.05; LDA score > 3.5). In all patients, multiple taxa differed significantly at baseline between short and long term survivors. Specifically, short term survivor fecal samples were significantly enriched in *porphyromonas, porphyromonadaceae, and dialister*, whereas long term survivor samples were significantly enriched in *Escherichia Shigella, Enterobacteriaceae, and Enterobacteriales* (P < 0.05; LDA score > 3.5, Fig. 2a,b). Our univariate analyses performed by Cox proportional hazard model identified *Pasteurellales, Haemophilus and Veillonella* as independent predictors for RFS and OS. We tested the relationship between these taxa and RFS and OS in our cohort by stratifying patients based on their relative abundance at baseline (Supplemental Fig. 2). We demonstrate that patients with high relative abundance of *Veillonella* at baseline showed a trend toward prolonged RFS and OS when compared to those with a low relative abundance at baseline (P = 0.08 and P = 0.054, respectively).

### Association between Gut Microbiota Profile and Immune Signatures

Because the gut microbiota is thought to influence disease progression partially through modulating local immune response, we analyzed the cervical tumors in our cohort of patients via flow cytometry on tumor brushings performed before week 1, week 3 and week 5 of radiation therapy. To identify features associated with high gut diversity, Spearman correlation analysis was conducted between immune signatures at each time point. High SDI was positively correlated with tumor infiltration of CD4 T cells at week 3, and CD4ki67+ T-cells at week 5, (Table 3 and Fig. 3a-d). These results suggest that patients with high gut diversity develop increased infiltration of activated CD4+ T-cell subsets.

**Table 3.** Correlation of baseline gut diversity (Inverse Shannon Diversity) with phenotype of tumor infiltrating lymphocytes during chemoradiation treatment. The percent of live lymphocytes expressing each marker was correlated with baseline Shannon diversity of the gut microbiome.

|  | P-value* | Q value | R<sub>2</sub> |
| --- | --- | --- | --- |
| CD4+Ki.67+ at T4 | 0.004‡ | 0.0714 |  |
| CD4+CD69+ at T3 | 0.004‡ | 0.1429 |  |
| CD4+PD1+ at T3 | 0.0367‡ | 0.2143 |  |
| CD4+CTLA4+ at T3 | 0.057 | 0.2857 |  |
| CD4+ |  |  | 0.1 |

\*P-value correlation of immune metric with baseline gut diversity
‡Significant *P* value.

**Fig 3.**
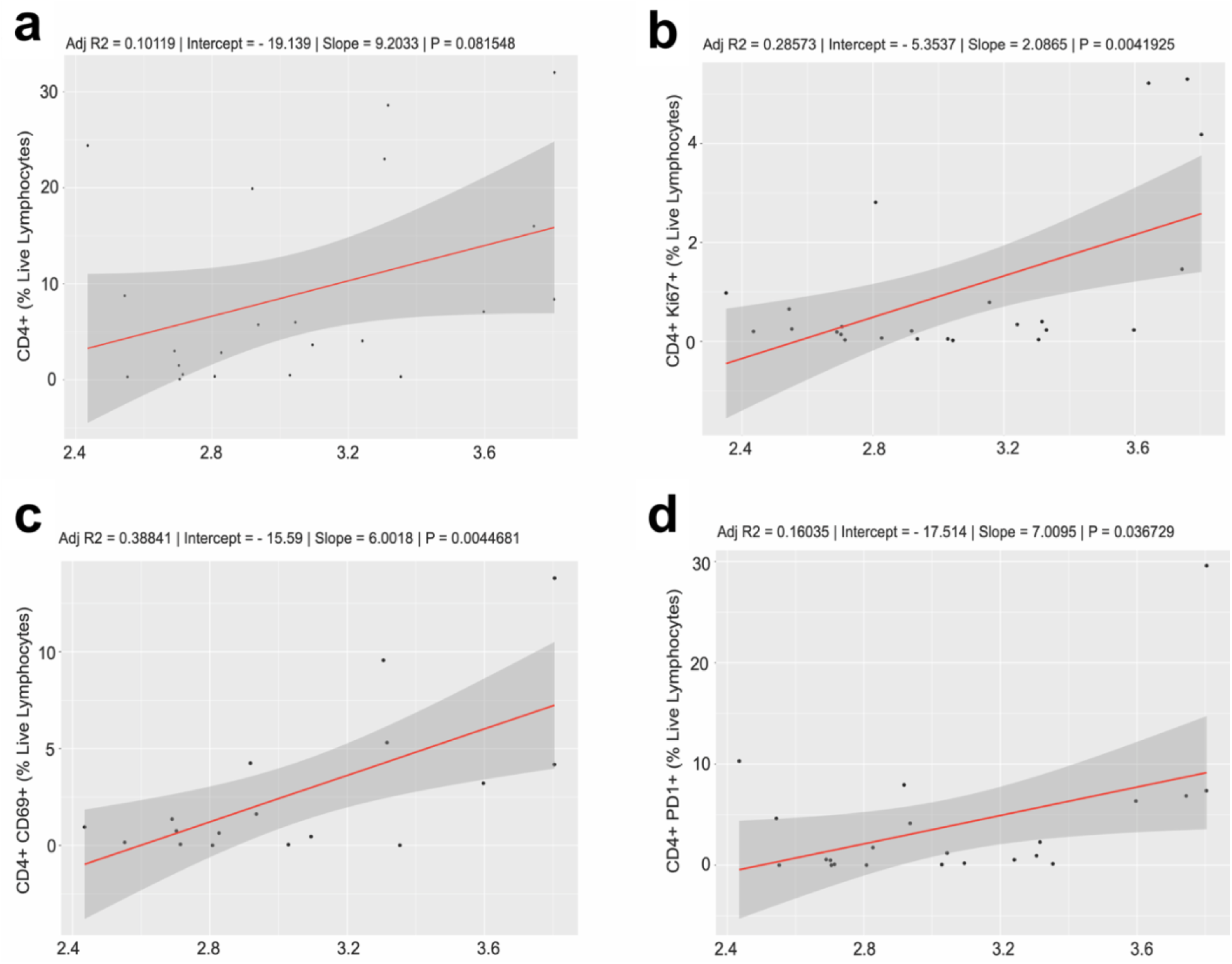
Correlation analysis of Shannon Diversity Index with tumor immune signatures. (A,B,C,D) Spearman correlations between Shannon Diversity Index and CD4+, CD4+ Ki67+, CD4+ CD69+, and CD4+ PD1+. Statistical analysis was performed by Spearman correlation or Mann-Whitney tests.

## DISCUSSION

The aim of this study was to identify independent gut microbial risk factors in cervical cancer patients receiving chemoradiation and to evaluate their impact on survival. We found BMI and gut diversity to be independent risk factors for RFS and OS in cervical cancer patients undergoing chemoradiation. Higher alpha gut diversity at baseline correlated with an improved RFS and OS. Our results indicate that overweight or obesity is a favorable prognostic factor independent of gut diversity. Additionally, our results demonstrate that patients with better clinical survival exhibit higher diversity as well as a distinct gut microbiome composition. Lastly, the association between gut diversity and local immune signatures highlights helper CD4+ T cells as potential mediators of antitumor immunity upon CRT treatment. Taken together, our results imply that the diversity of gut microbiota might be a shared benefit factor in those who respond well to CRT treatment.

It is now generally accepted that the gut microbiome modulates immune responses, antitumor immunity, and clinical outcomes in a variety of malignancies^9,11,19^. The gut microbiome is thought to affect both innate and adaptive immune responses. Specifically how the gut microbiome exerts its influence continues to be explored, but this explanation may have important implications if specific taxa are found to change host response to treatment via immunomodulation^6^. In our study, T helper cell profiles at baseline correlate with gut diversity. These results confer that T cells and response to CRT are likely affected by the gut microbiota independent of other factors such as BMI. Using multi-color flow cytometry we performed correlation analysis on individual immune signatures and microbiome diversity. The frequency of helper CD4+ T cells were chiefly identified. Cervical cancer is considered to be an immunogenic tumor because its origin is dependent on a persistent infection with human papillomavirus (HPV), most often HPV16 or HPV18^20^. Previous studies have reported that the number and functional orientation of tumor-infiltrating CD4+ and CD8+ T cells and the presence of M1 type macrophages strongly correlates with survival in patients with cervical cancer after chemoradiation^20,21^. T cells are capable of rapid antigen-specific responses and play critical roles in immune recall responses. In addition to the percentage of CD4+ T cell subsets, the increase in CD4 Ki67, CD4 CD69, and CD4 PD1 in patients with high microbiota diversity implies that the gut microbiome also modulates the proliferation of certain immune cell populations. Recent studies have already reported that chemoradiotherapy for cervical cancer induces unfavorable immune changes reflected by a decreased number of circulating lymphocytes, both CD4+ and CD8+ T cells, and an increased percentage in myeloid-cell populations, including myeloid-derived suppressor cells and monocytes^20^. Our study suggest that CD4+ T cells infiltrating the tumor microenvironment support and encourage the activity of other immune cells by releasing T cell cytokines.

We found gut diversity to be associated with a favorable response to CRT against cervical cancer. Considering the correlation between gut diversity and local helper T cells being reshaped upon CRT treatment, we propose that patients harboring a more diverse gut microbiota at baseline may benefit from CRT to a greater extent. This might be mediated by the reprogramming of local antitumor immune responses. The significance of our study lies in that the modulation of gut microbiota before treatment might provide an alternative way to enhance the efficacy of CRT, specifically in advanced staged disease in which systemic failure of current therapies represents a major challenge. Our results suggest that changes in the gut microenvironment contribute substantially to treatment success or failure, particularly in so-called immunogenic tumors like cervical cancer.

Our own group has previously characterized the gut microbiome of cervical cancer patients compared to healthy female controls, and have reported on differences in the relative abundance of specific taxa^22^. Our new findings support the hypothesis that organisms inhabiting the gut microbiome may be manipulated to improve cancer treatment response. Knowing specific gut microbial organisms that inhabit and undergo changes in patients with cervical cancer during CRT provides further insight into mechanisms that may modulate immune response and potentiate treatment outcomes in cancer patients. Researchers have already studied the treatment-enhancing utility of the gut microbiota in multiple areas of medicine^10,23^. Additionally, there is emerging data describing the influence of the gut microbiome as it pertains to radiotherapy^24^. Given that radiation can change the composition of the gut microbiome by altering the relative abundance of different taxa, we have to postulate whether radioresistant taxa ultimately alter the effectiveness of radiotherapy for cervical cancer^6,25,26^. The results of our study illustrate the potential of intentionally modifying the gut microbiota to accumulate CRT-tolerant species as an interventional strategy to enhance response of cervical cancer to CRT. Furthermore, determining whether changes in the human gut microbiome during CRT affect patients’ risk of treatment-related toxic effects may be an area that deserves further investigation.

The “obesity paradox”, which suggest a positive association between increasing BMI as it pertains to a specific disease, was firstly reported in heart failure^27^, but has since been described in a variety of disease processes, including other gynecologic cancers^28-30^. Theories centered around the “obesity paradox” suggest that patients with a high BMI may be better able to withstand cancer-induced consumption and stress compared with patients with a low BMI^31^. In uterine cancer it has been reported that the risk of recurrence differed significantly by BMI^32^. Specifically, a greater proportion of obese women met criteria for having a low risk of recurrence, while thin women tended to have a high-intermediate risk or recurrence. Many studies have investigated the impact of BMI on cervical cancer, but the association between weight and cervical cancer remains ambigous^33^. HPV is considered to be responsible for 99.7% of all cervical cancers^34^, however, it has been suggested that obesity may further increase this risk^35-37^. Other reports however do not report an association^38,39^. For example, a review by Lane *et al*. finally refuted the relationship between cervical cancer and obesity ultimately siting a lack of evidence^40^. The inconsistent conclusions among studies investigating the association between BMI and cervical cancer may be attributed to numerous factors including patient selection criteria, sample size and generalizability of the study population. Among these factors, patient selection criteria may be especially important, because tumor histology seems to be closely associated with BMI^37^.

The strengths of this study include the use of careful clinical staging, histopathology, and reliable phylogenetic and statistical analysis to assess bacterial community compositional changes using both microbial divergence and taxon-based methods. Additionally, we followed a complete protocol for 16S analysis ranging from the sample collection method, DNA extraction, and microbiome sequencing, thus limiting artifactual variations. Although this study has yielded intriguing findings, an important limitation is the small sample size. Consequently, the sample size limited our ability to weigh statistical power. Yet despite the relatively small size of this prospective study, large statistically significant differences were still observed, and we believe the results presented herein provide solid evidence elucidating the role of the gut microbiome as it pertains to the treatment of cervical cancer. We hope the integration of these data will produce actionable strategies geared toward targeting and manipulating the microbiome in order to ultimately improve cervical cancer therapy.

In conclusion, our study demonstrates that the diversity of gut microbiota is associated with a favorable response to chemoradiation. Additionally, compositional variation among patients correlated with short term and long-term survival. Our study demonstrates that gut diversity is a significant factor for predicting OS in CC patients undergoing CRT when BMI is accounted for, and may help explain the “obesity paradox” in cancer response. Moreover, analysis of immune cells from cervical tumor brush samples by flow cytometry revealed that an association between high microbial diversity, increased tumor infiltration of CD4+ lymphocytes and the activation of CD4 cells over the course of radiation therapy. The correlation between gut diversity and increased tumor infiltration of CD4+ lymphocytes suggest that patients harboring a more diverse gut microbiota at baseline may benefit from CRT to a greater extent. The significance of our study lies in that modulation of gut microbiota before chemoradiation might provide an alternative way to enhance treatment efficacy and improve treatment outcomes in cervical cancer patients. Additional studies exploring the relationship between gut diversity, chemoradiation, and treatment efficacy are needed to further understand the role of the gut microbiome in cervical cancer treatment.

## METHODS

### Participants and clinical data

Gut microbiome and cervical swab samples were collected prospectively from cervical cancer patients according to a protocol approved by The University of Texas MD Anderson Cancer Center Institutional Review Board (MDACC 2014-0543) for patients with biopsy-proven carcinoma of the cervix treated at MD Anderson and the Lyndon B. Johnson Hospital Oncology Clinic from September 22, 2015, to January 11, 2019. All patients had new diagnoses of locally advanced, nonmetastatic carcinoma of the cervix and underwent definitive CRT with EBRT followed by brachytherapy. Patients received a minimum of 45 Gy via EBRT in 25 fractions over 5 weeks with weekly cisplatin followed by two brachytherapy sessions at approximately weeks 5 and 7 with EBRT in between for gross nodal disease or persistent disease in the parametrium. Patients with stage IB1 cancer were given CRT due to the presence of nodal disease. Clinical variables, demographics, and pathologic reports were abstracted from electronic medical records.

### Sample collection and DNA extraction

Stool was collected from all patients by a clinician performing rectal exams at five time points (baseline; weeks 1, 3, and 5 of radiotherapy; and 3 months after CRT completion) using a matrix-designed quick-release Isohelix swab to characterize the diversity and composition of the microbiome over time. The swabs were stored in 20 μl of protease K and 400 μl of lysis buffer (Isohelix) and kept at −80°C within 1 h of sample collection.

### 16S rRNA gene sequencing and sequence data processing

16S rRNA sequencing was performed for fecal samples obtained from all patients at four time points to characterize the diversity and composition of the microbiome over time. 16S rRNA gene sequencing was done at the Alkek Center for Metagenomics and Microbiome Research at Baylor College of Medicine. 16S rRNA was sequenced using approaches adapted from those used for the Human Microbiome Project^41^. The 16S rDNA V4 region was amplified via polymerase chain reaction with primers that contained sequencing adapters and single-end barcodes, allowing for pooling and direct sequencing of polymerase chain reaction products. Amplicons were sequenced on the MiSeq platform (Illumina) using the 2 × 250-bp paired-end protocol, yielding paired-end reads that overlapped nearly completely. Sequence reads were demultiplexed, quality-filtered, and subsequently merged using the USEARCH sequence analysis tool (version 7.0.1090) (4). 16S rRNA gene sequences were bundled into operational taxonomic units at a similarity cutoff value of 97% using the UPARSE algorithm^42^. To generate taxonomies, operational taxonomic units were mapped to an enhanced version of the SILVA rRNA database containing the 16Sv4 region. A custom script was used to create an operational taxonomic unit table from the output files generated as described above for downstream analyses of α-diversity, β-diversity, and phylogenetic trends. Principal coordinates analysis was performed by institution and sample set to make certain no batch effects were present.

### Flow Cytometry

Immunostaining was performed according to standard protocols^43^. Cells were fixed using the Foxp3/Transcription Factor Staining Buffer Set (eBioscience, Waltham, MA) and stained with a 16 color panel with antibodies from Biolegend (San Diego, CA), BD Bioscience (San Jose, CA), eBioscience (Waltham, MA), and Life Technologies (Carlsbad, CA). Analysis was performed on a 5-laser, 18 color LSRFortessa X-20 Flow Cytometer (BD Biosciences, San Jose, CA). Analysis was performed using FlowJo version 10 (Flowjo LLC, Ashland, OR). We then followed a similar previously published method^43^. Briefly, the cells were incubated with the antibodies for surface markers at 4°C in dark for 30 minutes. They were then washed twice with FACS buffer and fixed and permeabilized with FOXP3 Fix/perm Kit (ThermoFisher Scientific, Waltham, MA). Next, intracellular staining was performed by preparing the antibodies in permeabilization buffer and incubating the cells for 30 minutes at 4°C in the dark. Cells were washed with FACS buffer twice and prepared for acquisition on an LSR Fortessa X-20 analyzer at the Flow Cytometry Core at MD Anderson Cancer Center and were analyzed using FlowJo software (FlowJo, LLC, Ashland, OR). Compensation controls were prepared using OneComp ebeads (eBioscience, Waltham, MA) and fluorescence minus one controls were used^43^.

### Statistical analyses

For microbiome analysis, rarefaction depth was set at 7066 reads. The Shannon diversity index (SDI) was used to evaluate α-diversity (within samples), and principle coordinates analysis of unweighted UniFrac distances was used to examine β-diversity (between samples). Patient and tumor characteristics were analyzed by univariate and multivariate Cox regression models for Recurrence-free survival (RFS) and Overall survival (OS) based on univariate p-value < 0.1. Characteristics included age, body mass index (BMI), race, FIGO stage, grade, histology, nodal status, smoking status, antibiotic use and max tumor size. For each outcome of interest, a multivariate Cox regression analysis was performed to adjust for the effects of prognostic factors identified on univariate analysis as influencing survival in cervical cancer. These analyses were conducted using covariates with p ≤ 0.1 in a stepwise fashion. We also ran a correlation analysis of alpha diversity metrics with tumor flow cytometry markers using a linear regression and Spearman’s correlation. Alpha (within sample) diversity was evaluated using SDI. The relative abundance of microbial taxa, classes, and genera present in long term vs short term survivors was determined using LDA Effect Size^44^, applying the one-against-all strategy with a threshold of 3.5 for the logarithmic LDA score for discriminative features and α of 0.05 for factorial Kruskal-Wallis testing among classes. Long term survivors were classified as patients who had a follow up of two years or more and were alive at time of last follow up, while short term survivors had a follow up of one year or less. LDA Effect Size analysis was restricted to bacteria present in 20% or more of the study population. Kaplan-Meier curves were generated for patients with normal BMI and overweight/obese BMI based on Cox analysis and clostridia abundance. The significance of differences was determined using the log-rank test. Statistical significance was set at an alpha of 5% for a two-sided p-value. Analyses were conducted using Rstudio version Orange Blossom – 1.2.5033.

## Data Availability

N/A

## ACKNOWLEDGEMENTS

This research was supported in part by the Radiological Society of North America Resident/Fellow Award (to L.E.C.), the National Institutes of Health (NIH) through MD Anderson’s Cancer Center Support Grant P30CA016672, the Emerson Collective and the National Institutes of Health T32 grant #5T32 CA101642-14 (T.T.S). This study was partially funded by The University of Texas MD Anderson Cancer Center HPV-related Cancers Moonshot (L.E.C and A.K.). The human subjects who participated in this study are gratefully acknowledged.

## AUTHOR CONTRIBUTION

All authors were involved in subject identification, data collection, interpretation of the statistical analysis, and review and approval of the final manuscript. The study concept was conceived by L.E.C., A.K., M.B.E.A, and T.T.S. The manuscript was written by T.T.S.

---

**Supplemental Fig 1.**
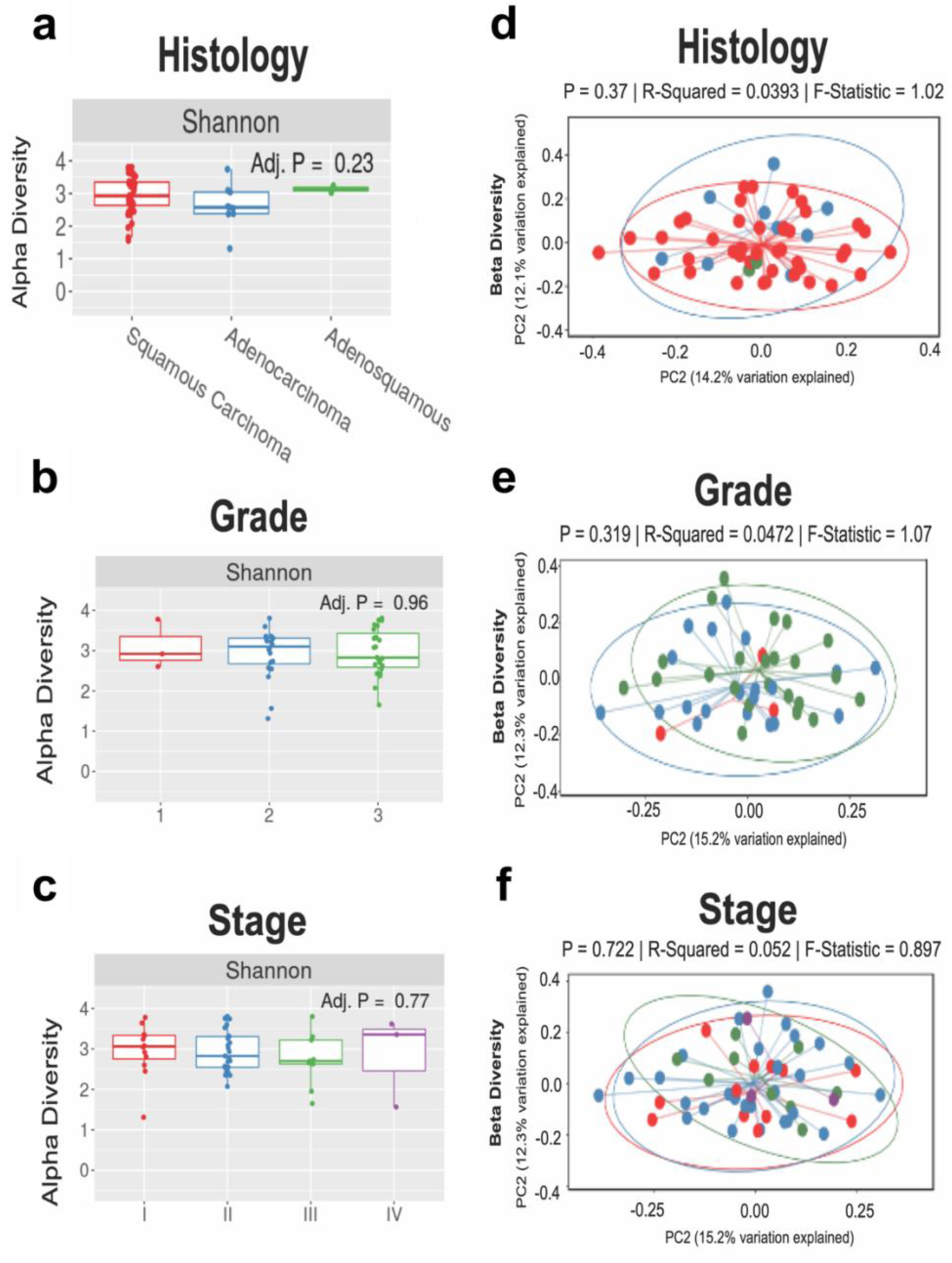
The fecal microbiota of individuals with cervical cancer. The fecal microbiota of individuals with cervical cancer by demographics. Diversity (within sample diversity) was measured using the Shannon diversity metric and Beta diversity (between sample diversity) was determined by unweighted Unifrac. No differences were observed in either metric by cancer histology (A,D), grade (B,E) or cancer stage (C,F).

**Supplemental Fig 2.**
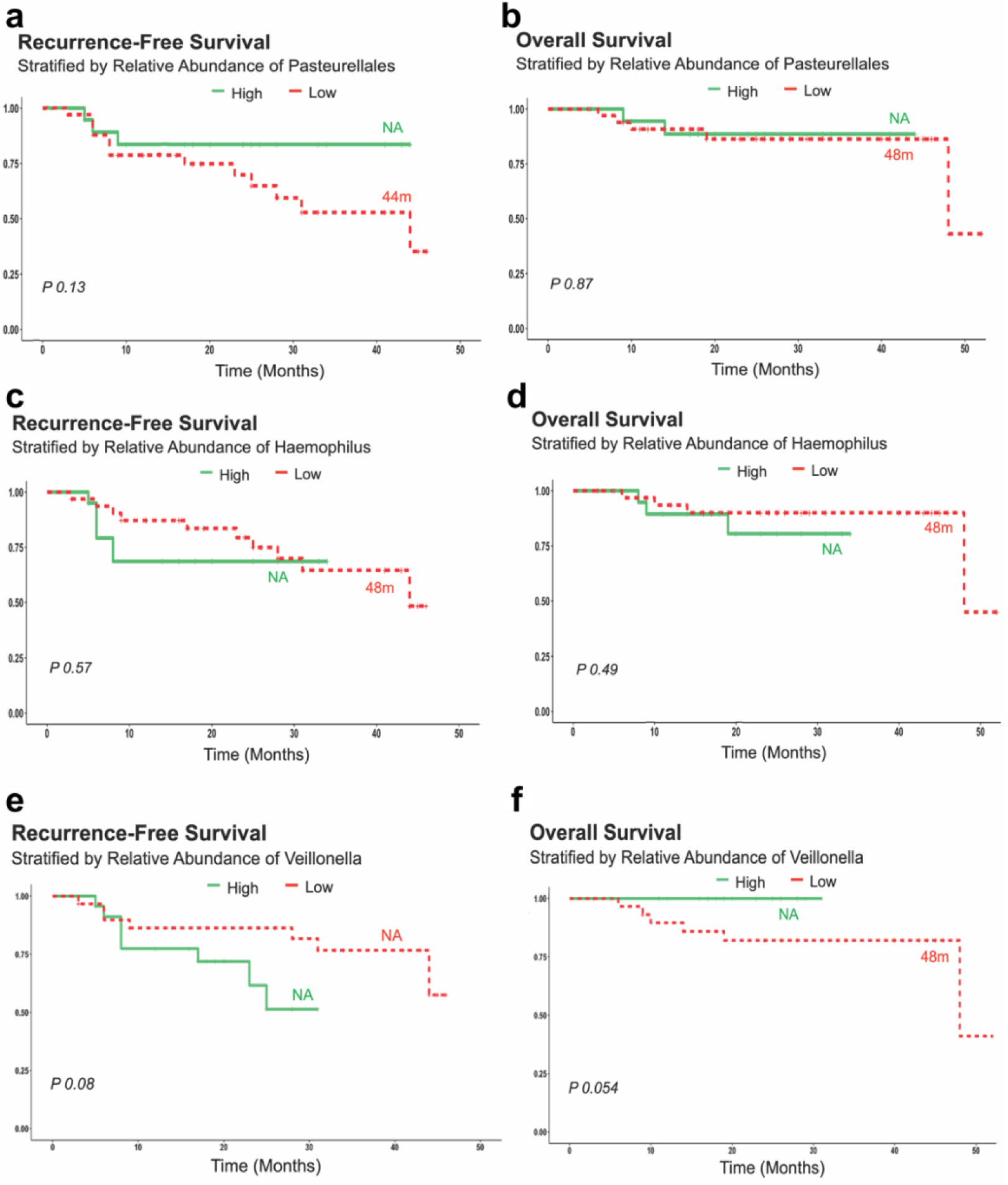
Relationship between gut diversity and BMI. Kaplan-Meier curves for (A) recurrence free survival, (B) overall survival stratified by relative abundance of *Pasteurellales*. Kaplan-Meier curves for (C) recurrence free survival, (D) overall survival stratified by relative abundance of *Haemophilus*. Kaplan-Meier curves for (E) recurrence free survival, (F) overall survival stratified by relative abundance of *Veillonella*. Cases represent patients.

**Supplemental Table 1.** Patient and tumor characteristics (N=55)

|  | N | (%) |
| --- | --- | --- |
| **Median age, yrs (range)** | 48 (28-72) | — |
| **BMI, Mean (SD), kg/m2** | 28.7(6.06) | — |
| **Race/Ethnicity** |  |  |
| Asian | 2 | (36.4) |
| Black | 4 | (18.2) |
| Hispanic | 24 | (43.6) |
| White | 24 | (43.6) |
| Other | 1 | (1.8) |
| **FIGO Stage** |  |  |
| IA1 | 1 | (1.8) |
| IA2 | 0 | (0) |
| IB1 | 5 | (9.09) |
| IB2 | 6 | (10.9) |
| IIA | 3 | (5.45) |
| IIB | 28 | (50.9) |
| IIIA | 9 | (16.3) |
| IIIB | 0 | (0) |
| IVA | 3 | (5.45) |
| IVB | 0 | (0) |
| **Grade** |  |  |
| Well | 4 | (7.2) |
| Moderate | 20 | (36.3) |
| Poor | 25 | (45.4) |
| Unknown | 6 | (10.9) |
| **Histology** |  |  |
| Squamous | 43 | (78.1) |
| Adenocarcinoma | 8 | (18.1) |
| Adenosquamous | 3 | (3.63) |
| **Node Level on PET** |  |  |
| Common Iliac | 9 | (16.3) |
| External Iliac | 23 | (41.8) |
| Internal Iliac | 5 | (9.09) |
| Para-Aortic | 3 | (5.45) |
| None | 15 | (27.2) |
| **Median cervical tumor size (cm)** | 5.4 | — |
| **Smoking status** |  |  |
| Current | 4 | (7.27) |
| Former | 20 | (36.3) |
| Never | 31 | (56.3) |
| **Antibiotic Use** |  |  |
| No | 5 | (9.1) |
| Yes | 50 | (90.9) |
| **Brachytherapy** |  |  |
| HDR | 21 | (38.2) |
| PDR | 34 | (61.8) |
| **Concurrent Chemotherapy (cycles)** |  |  |
| *Cisplatin* |  |  |
| (1-3) | 2 | (3.6) |
| (≥4) | 51 | (92.7) |
| *Carboplatin* |  |  |
| (2) | 1 | (1.8) |
| *Carboplatin + Cisplatin* |  |  |
| (2)+(2) | 1 | (1.8) |

FIGO- International Federation of Gynecology and Obstetrics
HDR-High Dose Rate
PDR- Pulsed Dose Rate

**Supplemental Table 2.** Univariate Cox regression analysis for recurrence-free survival – Alpha Diversity all time points.

| **Characteristics** | **Univariate model** |  |
| --- | --- | --- |
|  | **HR (95% CI)** | ***P* value** |
| **Observed OTU** |  |  |
| Baseline | 0.99 (0.97-1) | 0.21 |
| Week 1 | 1 (0.97-1) | 0.69 |
| Week 3 | 0.99 (0.96-1) | 0.59 |
| Week 5 | 1 (0.98-1) | 0.71 |
| Week 12 | 1 (0.98-1) | 0.77 |
| **Shannon** |  |  |
| Baseline | 0.51 (0.23-1.1) | 0.087 |
| Week 1 | 0.94 (0.2-4.4) | 0.94 |
| Week 3 | 1.2 (0.25-5.6) | 0.83 |
| Week 5 | 0.83 (0.35-1.9) | 0.66 |
| Week 12 | 2.7 (0.13-57) | 0.51 |
| **Simpson** |  |  |
| Baseline | 0.025 (0.00036-1.7) | 0.087 |
| Week 1 | 13 (1.4e-05-1.2e+07) | 0.13 |
| Week 3 | 52 (6.6e-05-4.1e+07) | 0.57 |
| Week 5 | 0.31 (0.013-7.8) | 0.48 |
| Week 12 | 130000 (7.5e-13-2.2e+22) | 0.56 |

| Characteristics | Univariate model |  |
| --- | --- | --- |
|  | HR (95% CI) | *P* value |
| **Inverse Simpson** |  |  |
| Baseline | 0.93 (0.84-1) | 0.11 |
| Week 1 | 0.96 (0.79-1.2) | 0.69 |
| Week 3 | 1 (0.95-1.1) | 0.34 |
| Week 5 | 1 (0.92-1.2) | 0.54 |
| Week 12 | 1.1 (0.81-1.4) | 0.59 |
| **Fisher** |  |  |
| Baseline | 0.95 (0.88-1) | 0.23 |
| Week 1 | 0.97 (0.86-1.1) | 0.66 |
| Week 3 | 0.96 (0.83-1.1) | 0.6 |
| Week 5 | 1 (0.91-1.2) | 0.69 |
| Week 12 | 1 (0.89-1.2) | 0.81 |

*CI*, Confidence interval; *HR*, hazard ratio.
\*Significant hazard ratios.
‡Significant *P* value.

**Supplemental Table 3.** Univariate Cox regression analysis for overall survival – Alpha Diversity all time points.

| Characteristics | Univariate model |  |
| --- | --- | --- |
|  | HR (95% CI) | *P* value |
| **Observed OTU** |  |  |
| Baseline | 0.98 (0.95-1) | 0.14 |
| Week 1 | 0.98 (0.94-1) | 0.35 |
| Week 3 | 0.97 (0.92-1) | 0.21 |
| Week 5 | 1 (0.96-1) | 0.98 |
| Week 12 | NA (NA-NA) | 1 |
| **Shannon** |  |  |
| Baseline | 0.34 (0.1-1.1) | 0.08 |
| Week 1 | 0.48 (0.063-3.7) | 0.48 |
| Week 3 | 1.2 (0.25-5.6) | 0.83 |
| Week 5 | 0.23 (0.037-1.4) | 0.11 |
| Week 12 | NA (NA-NA) | 1 |
| **Simpson** |  |  |
| Baseline | 0.0059 (1.2e-05-2.9) | 0.1 |
| Week 1 | 0.45 (1.1e-08-1.8e+07) | 0.93 |
| Week 3 | 0.009 (1.7e-07-490) | 0.4 |
| Week 5 | 1.4 (0.00063-3200) | 0.93 |
| Week 12 | NA (NA-NA) | 1 |
|  | HR (95% CI) | *P* value |
| **Inverse Simpson** |  |  |
| Baseline | 0.85 (0.7-1) | 0.13 |
| Week 1 | 0.86 (0.62-1.2) | 0.39 |
| Week 3 | 0.81 (0.61-1.1) | 0.15 |
| Week 5 | 0.89 (0.66-1.2) | 0.46 |
| Week 12 | NA (NA-NA) | 1 |
| **Fisher** |  |  |
| Baseline | 0.91 (0.79-1) | 0.15 |
| Week 1 | 0.91 (0.74-1.1) | 0.34 |
| Week 3 | 0.84 (0.64-1.1) | 0.22 |
| Week 5 | 0.99 (0.79-1.3) | 0.94 |
| Week 12 | NA (NA-NA) | 1 |

*CI*, Confidence interval; *HR*, hazard ratio.
\*Significant hazard ratios.
‡Significant *P* value.

